# Procedural and Clinical Outcomes of High-Frequency-Low-Tidal-Volume Ventilation Plus Rapid-Atrial Pacing in Paroxysmal Atrial Fibrillation Ablation

**DOI:** 10.1101/2024.08.12.24311903

**Authors:** Paul C. Zei, Joan Rodriguez-Taveras, Daniela Hincapie, Jose Osorio, Isabella Alviz, Andres F. Miranda-Arboleda, Mohamed Gabr, Christopher Thorne, Joshua R. Silverstein, Amit J. Thosani, Allyson L. Varley, Fernando Moreno, Daniel A. Zapata, Benjamin D’Souza, Anil Rajendra, Saumil Oza, Linda Justice, Ana Baranowski, Huy Phan, Alejandro Velasco, Charles C. Te, Matthew C. Sackett, Matthew J. Singleton, Anthony R. Magnano, David Singh, Richard Kuk, Nathaniel A. Steiger, William H. Sauer, Jorge E. Romero

**Affiliations:** Cardiac Arrhythmia Service, Division of Cardiovascular Medicine, Brigham and Women’s Hospital, Harvard Medical School, Boston, MA, USA; Department of Medicine, Boston Medical Center & Boston University Chobanian & Avedisian School of Medicine, Boston, MA, USA; HCA Florida, Miami, FL, USA; Heart Rhythm Clinical Research Solutions, Birmingham, AL, USA; Allegheny Health Network, Baden, PA, USA; Allegheny Health Network, Pittsburgh, PA, USA; Perelman School of Medicine at the University of Pennsylvania, Philadelphia, PA, USA; Arrhythmia Institute, Grandview Medical Group, Birmingham, AL, USA; Ascension Medical Group, St Vincent’s Cardiology, Jacksonville, FL, USA; Valley Heart Rhythm Specialists, PLLC, Chandler, AZ, USA; University of Texas Health San Antonio, San Antonio, TX, USA; Oklahoma Heart Hospital, Oklahoma City, OK, USA; Centra Heart and Vascular Institute, Lynchburg, VA, USA; WellSpan Health, York, PA, USA; Queen’s Heart Institute, Honolulu, Hawaii, USA

**Keywords:** Paroxysmal atrial fibrillation, High-frequency low-tidal volume ventilation, Pulmonary vein isolation, Radiofrequency catheter ablation, Rapid atrial pacing

## Abstract

**Background:** High-frequency low-tidal volume (HFLTV) ventilation is a safe and cost-effective strategy that improves catheter stability, first-pass pulmonary vein isolation, and freedom from all-atrial arrhythmias during radiofrequency catheter ablation (RFCA) of paroxysmal atrial fibrillation (PAF). However, the incremental value of adding rapid-atrial pacing (RAP) to HFLTV-ventilation has not yet been determined.

**Objective:** To evaluate the effect of HFLTV-ventilation plus RAP during RFCA of PAF on procedural and long-term clinical outcomes compared to HFLTV-ventilation alone.

**Methods:** Patients from the REAL-AF prospective multicenter registry, who underwent RFCA of PAF using either HFLTV+RAP or HFLTV ventilation alone from April 2020 to February 2023 were included. The primary outcome was freedom from all-atrial arrhythmias at 12-months. Secondary outcomes included procedural characteristics, long-term clinical outcomes, and complications.

**Results:** A total of 545 patients were included (HFLTV+RAP=327 vs. HFLTV=218). There were no significant differences in baseline characteristics. No differences in procedural (HFLTV+RAP 74 [57-98] vs. HFLTV 66 [53-85.75] min, p=0.617) and RF (HFLTV+RAP 15.15 [11.22-21.22] vs. HFLTV 13.99 [11.04-17.13] min, p=0.620) times. Both groups had a similar freedom from all-atrial arrhythmias at 12 months (HFLTV+RAP 82.68% vs. HFLTV 86.52%, HR=1.43, 95% CI [0.94-2.16], p=0.093). There were no differences in freedom from AF-related symptoms (HFLTV+RAP 91.4% vs. HFLTV 93.1%, p=0.476) and rate of AF-related hospitalizations (HFLTV+RAP 1.5% vs. HFLTV 2.8%, p=0.320) between groups. Procedure-related complications were low in both groups (HFLTV+RAP 0.6% vs. HFLTV 0%, p=0.247).

**Conclusion:** In patients undergoing RFCA for PAF, adding RAP to HFLTV-ventilation was not associated with improved procedural and long-term clinical outcomes.

**Condensed Abstract:** High-frequency low-tidal volume (HFLTV) ventilation is a safe and cost-effective strategy that improves the efficiency and efficacy of radiofrequency catheter ablation (RFCA) for paroxysmal atrial fibrillation (PAF). Our study aimed to assess the effect of HFLTV-ventilation plus rapid-atrial pacing (RAP) during RFCA of PAF on procedural and long-term clinical outcomes compared to HFLTV-ventilation alone. Patients from the REAL-AF registry who underwent PAF-RFCA from April 2020-February 2023, using HFLTV-ventilation alone or in combination with RAP, were analyzed. Our study concluded that in patients undergoing RFCA for PAF, adding RAP to HFLTV-ventilation was not associated with improved procedural and long-term clinical outcomes.

## Introduction

Atrial fibrillation (AF) is the most common sustained arrhythmia in adults, affecting over 50 million individuals worldwide. Its incidence and prevalence in the United States have increased progressively, leading to significant morbidity and mortality (1,2). Recent guidelines emphasize the superiority of catheter ablation over medical therapy with antiarrhythmic drugs (AADs) for the treatment of paroxysmal and persistent AF (PAF and PeAF) (3,4). Several randomized controlled trials (RCTs) have shown significant advantages of catheter ablation as first-line therapy for AF in improving freedom from all-atrial arrhythmias at 1-year follow-up (5-10). However, post-procedure long-term clinical outcomes remain suboptimal, with a previous RCT demonstrating AF recurrence rates as high as 35% and pulmonary vein (PV) reconnections in up to 52% of AF recurrence cases (11). Therefore, achieving durable PV isolation (PVI) is of paramount importance for catheter ablation in PAF, since the electrical reconnection of the PVs has been associated with early and long-term AF recurrence (11,12). Over the past decade, efforts have been made to improve outcomes in radiofrequency catheter ablation (RFCA) of AF, including the development of catheter irrigation systems and contact force (CF) sensing catheters (13), as well as the optimization of procedural techniques with the utilization of high-power short-duration (HPSD) ablation (14). Maintaining stable CF throughout radiofrequency (RF) application improves lesion size and promotes lesion transmurality, thereby enhancing procedural and clinical outcomes (15-17). Nevertheless, ensuring optimal catheter-tissue contact during RFCA represents a significant challenge, primarily due to the impact of external factors such as cardiac contraction and diaphragmatic excursion on catheter stability and CF (18). General anesthesia (GA) has been demonstrated to improve outcomes in RFCA of AF by minimizing patient movement (19,20). Nonetheless, its efficacy is limited by CF variations associated with the high tidal volumes used in standard ventilation (SV) (19). Consequently, alternative mechanical ventilation techniques, such as high-frequency jet ventilation (HFJV) (21,22), and high-frequency low-tidal-volume ventilation (23-25), have been developed. These techniques have been shown to improve procedural and clinical outcomes by reducing diaphragmatic excursion, thereby enhancing catheter stability, lesion transmurality, and lesion durability. Rapid atrial pacing (RAP) alone (26) or in combination with HFJV (21) has been shown to reduce CF variability, thereby enhancing catheter stability and lesion formation by inducing heart rate acceleration (26). This strategy can potentially reduce cardiac contraction, maximize catheter stability, and improve procedural characteristics and long-term clinical outcomes. RAP may also elevate the rate of electroanatomic data acquisition, theoretically decreasing mapping time. However, the incremental value of adding RAP to HFLTV ventilation on procedural and long-term clinical outcomes remains uncertain, particularly given the need to introduce a reference catheter, typically in the coronary sinus (CS), which may require an additional vascular access site. The objective of this multicenter prospective study was to evaluate the effect of HFLTV ventilation plus RAP during RFCA of PAF on procedural and long-term clinical outcomes compared to HFLTV ventilation alone.

## Methods

### Study Design and Participants

This prospective multicenter cohort study from the REAL-AF Registry (Real-World Experience of Catheter Ablation for the Treatment of Paroxysmal and Persistent Atrial Fibrillation, NCT04088071) included patients who underwent RFCA for symptomatic PAF using either HFLTV ventilation plus RAP (through a Decapolar CS catheter or multispline catheter in the superior vena cava (SVC)) or HFLTV ventilation alone. The study period spanned from April 2020 to February 2023 across 16 centers in the United States. Detailed descriptions of the registry design and objectives have been published previously (27). The study had a minimum follow-up period of 12 months. Comprehensive clinical and procedural data were systematically and prospectively collected during the initial ablation procedure, utilizing standardized case report forms (CRFs) and an Electronic Data Capture (EDC) system via the 3PHCloud platform to enable for a double-step verification process for the information of each enrolled patient. Periprocedural management followed the standard practice of each operator and center. This analysis included patients who had completed 12 months of follow-up, which encompassed clinical visits (at 3 and 12 months) and long-term ambulatory monitoring (at 6 and 12 months). Patients with PeAF and procedures that were performed with SV or any ventilation strategy other than HFLTV ventilation were excluded from the analysis.

### Ventilation Protocol

The HFLTV ventilation protocol was conducted by the anesthesia team. The respiratory rate was set to approximately 25-30 breaths per minute with a tidal volume of 200-250 ml (3-3.5 mL/kg), aiming for a minute ventilation of 6 L/min. These ventilatory settings were only used during RF time to minimize the potential risk of CO_2_ retention while preserving an end-tidal CO_2_ level below 50 mmHg. Hyperventilation periods were used as needed to achieve this target as “recruitment breaths”.

### Ablation Procedure

All individuals underwent RFCA for PAF under general anesthesia with either uninterrupted oral anticoagulation (OAC) or minimally interrupted OAC (apixaban held on the morning of the procedure). Operators had the autonomy to decide whether to use ultrasound-guided venous access. Following transseptal access, a multipolar mapping catheter chosen by the operator (PentaRay^TM^ or OctaRay^TM^, Biosense Webster, Inc., CA, USA) was used to create a three-dimensional shell and conduct comprehensive voltage mapping of the left atrium and the PVs. Procedures were performed using the ThermoCool SmartTouch SurroundFlow® CF-sensing catheter (STSF; Biosense Webster, Inc., CA, USA) and conducted with zero to minimal fluoroscopy use. PVI was performed using high-power short-duration (HPSD) ablation (40-50W, <20 seconds per lesion) (28) with wide-area circumferential ablation. Impedance drop, CF, and ablation index were monitored in real-time. All RF lesions were guided by the lesion tag assignment software VISITAG SURPOINT® Module (Biosense Webster, Inc., CA, USA), with a minimum location stability requirement of 2.5 mm for 4 seconds (29). The ablation procedures were typically guided by a standardized ablation index approach, with 4-5 mm distance between each lesion. The CF goal for each ablation lesion was prespecified to be between 10 and 20 g. Esophageal temperature monitoring and/or cooling devices were used for all patients, and ablation was stopped if esophageal temperature increased by ≥1°C. After completion of the ablation procedure, all PVs were assessed for bidirectional block (entrance and exit block). Vascular closure devices (Perclose ProGlide™ SMC System [Abbott] or VASCADE MVP® [Cardiva Medical]) or purse-string suture were selected based on operator’s preference. Cavotricuspid isthmus ablation was performed if a history of typical atrial flutter (AFL) was documented.

### Atrial Pacing Protocol

RAP was performed based on physician workflow using either a catheter placed in the CS (Decapolar Mapping Catheter-Decanav, Biosense Webster, Inc., CA, USA) or a multispline catheter in the SVC (PentaRay^TM^ or OctaRay^TM^, Biosense Webster, Inc., CA, USA). If a patient was found to be in AF during the procedure, direct current biphasic cardioversion was conducted to restore normal sinus rhythm prior to performing RAP. The atrium was initially paced at 500-600 milliseconds based on patient’s blood pressure tolerability and 1:1 atrioventricular (AV) conduction. If there was a lack of 1:1 AV conduction while pacing at 500 milliseconds, the pacing rate was adjusted accordingly and reduced to 550, or 600 milliseconds as needed to promote 1:1 AV conduction. These data were collected at the physician level via survey and then validated prior to analysis (30).

### Patient Follow-Up

Participants were monitored for a minimum of 12 months post-ablation procedure, with clinical assessments conducted at 3 and 12 months. Each assessment included clinical examination and electrocardiographic evaluation with a 12-lead electrocardiogram (ECG) at every visit. Also, continuous rhythm monitoring was conducted at 6 and 12 months of follow-up. The methods and duration of follow-up cardiac monitoring were conducted according to each center’s standard practice. Arrhythmia recurrence was also tracked using preexisting implantable cardiac devices, implantable loop recorders (ILRs), or twice-daily ECG checks using the Kardia Mobile device (AliveCor). Additional cardiac event monitors or ILRs were placed if AF-related symptoms were reported in the follow-up visits to address for symptomatic AF recurrence.

### Study Outcomes

The primary outcome was freedom from all-atrial arrhythmias at 12 months of follow-up. Arrhythmia recurrence was defined as any episode of AF, AFL, or atrial tachycardia (AT) lasting ≥30 seconds, and confirmed by a 12-lead ECG, rhythm strip, device electrograms, or mobile cardiac telemetry, reported after a 90-day blanking period following the ablation procedure and until the end of the 12-month follow-up. Secondary outcomes included freedom from each subtype of atrial arrhythmia (AF, AFL, AT) at the 12-month follow-up, procedural characteristics including procedural time, RF time, first-pass PVI, long-term clinical outcomes (freedom from AF-related symptoms, rate of AF-related hospitalizations) and acute and long-term procedure-related complications (e.g., ischemic stroke, transient ischemic attack [TIA], pericardial effusion/cardiac tamponade, phrenic nerve injury, atrioesophageal fistula or vascular access complications such as groin hematoma).

### Statistical Analysis

Categorical variables were presented as absolute numbers and percentages. The Shapiro-Wilk test was used to assess the normality of data distribution. Continuous data were expressed as mean ± standard deviation (SD) for normally distributed data or as median with interquartile range (IQR) for non-normally distributed data. Student’s T-test and Mann-Whitney U test were used for univariate comparisons depending on data distribution. The Chi-square (*X^2^*) test or Fisher’s exact test was used for categorical variables. An adjusted multivariate analysis to control for operator as a random effect was done for the comparison of procedural characteristics between the groups. Freedom from all-atrial arrhythmias at 12 months of follow-up was depicted using a time-to-event plot with the Kaplan-Meier method. Differences between the groups were calculated using the Cox Proportional Hazards model, adjusting for the procedure operator as a random effect. Prior to these analyses, the Schoenfeld test was conducted to ensure that the proportional hazard assumption was not violated. Patients who were lost to follow-up were excluded from the statistical analysis, and patients with missing data were censored. Two-tailed *p*-values <0.05 were considered to indicate statistical significance. Statistical analyses were performed using SPSS Statistics Software for Windows Version 26.0 (IBM Corporation, Armonk, New York), and R Studio.

### Institutional Review Board Approval

The REAL-AF registry was approved by the Western Institutional Review Board–Copernicus Group on October 31, 2018. Furthermore, institutional review boards of each participating institution approved the participation of enrolling centers adhering to the principles outlined in the Declaration of Helsinki.

## Results

### Baseline Characteristics

A total of 545 patients were included (mean age of 65.89 ± 10.42 years; 53.5% male), 327 patients underwent RFCA for PAF with both HFLTV ventilation plus RAP, and 218 patients underwent RFCA under HFLTV ventilation protocol alone (**Figure 1**). There were no statistically significant differences in baseline characteristics between groups. Both groups had a similar distribution of comorbidities, such as hypertension (HFLTV+RAP 65.7% vs. HFLTV 68.3%, p=0.528), diabetes mellitus (HFLTV+RAP 15% vs. HFLTV 12.8%, p=0.482) and vascular disease (HFLTV+RAP 7.1% vs. HFLTV 10.1%, p=0.212). A minority of patients had a past medical history of stroke (HFLTV+RAP 7.1% vs. HFLTV 9.9%, p=0.235). There was no difference in the rate of AADs use before the procedure (HFLTV+RAP 44.5% vs. HFLTV 41.5%, p=0.489). There were no differences in echocardiographic parameters within the groups, with comparable left ventricular ejection fraction (LVEF) (HFLTV+RAP 56.56 ± 8.55% vs. HFLTV 58.51 ± 7.89%, p=0.055) and left atrial volume index (HFLTV+RAP 25.1 ± 10.14 mL/m2 vs. HFLTV 29.09 ± 9.15 mL/m2, p=0.209) (**Table 1**).

**Figure 1.**
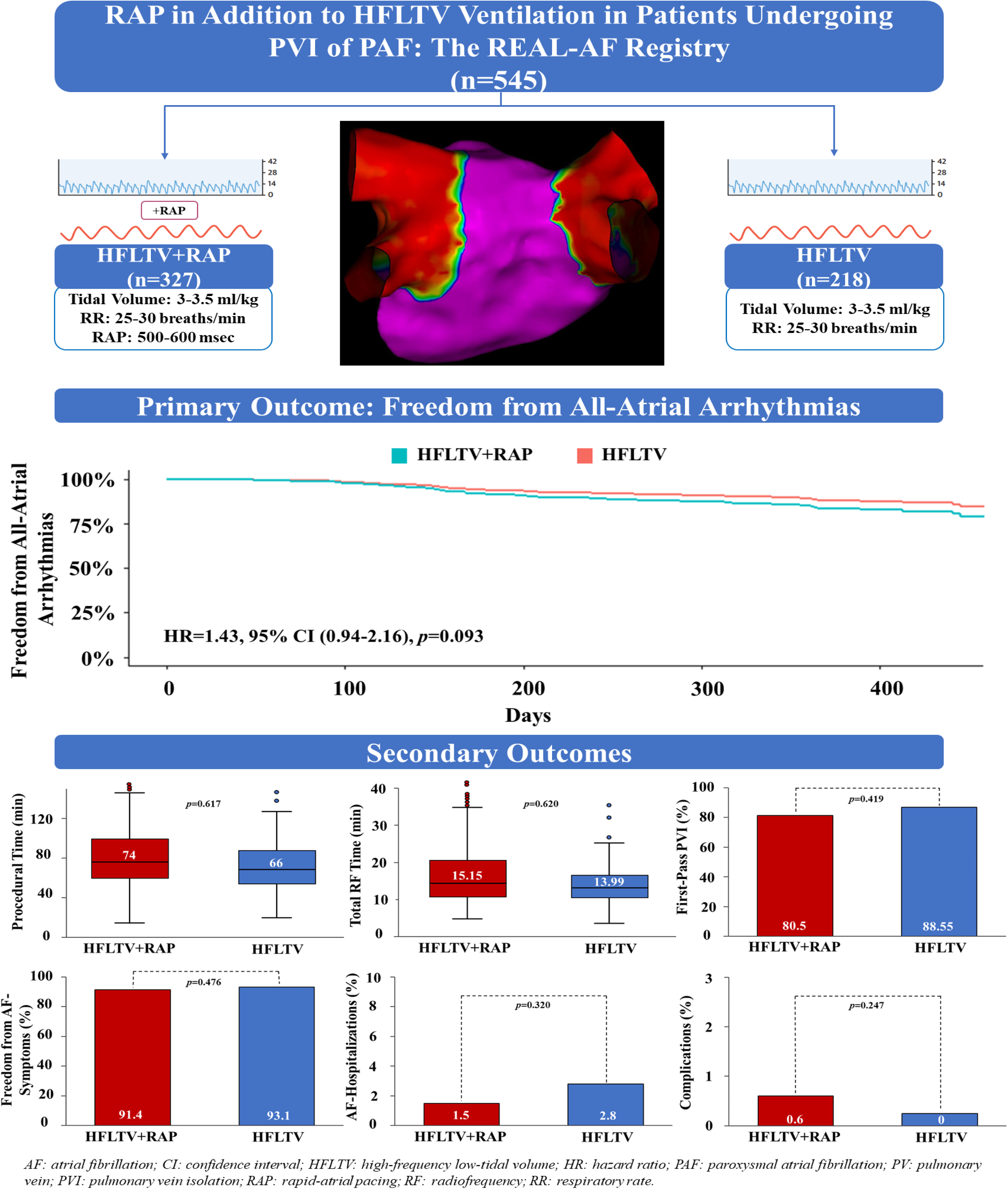
Study Design.

**Table 1.**
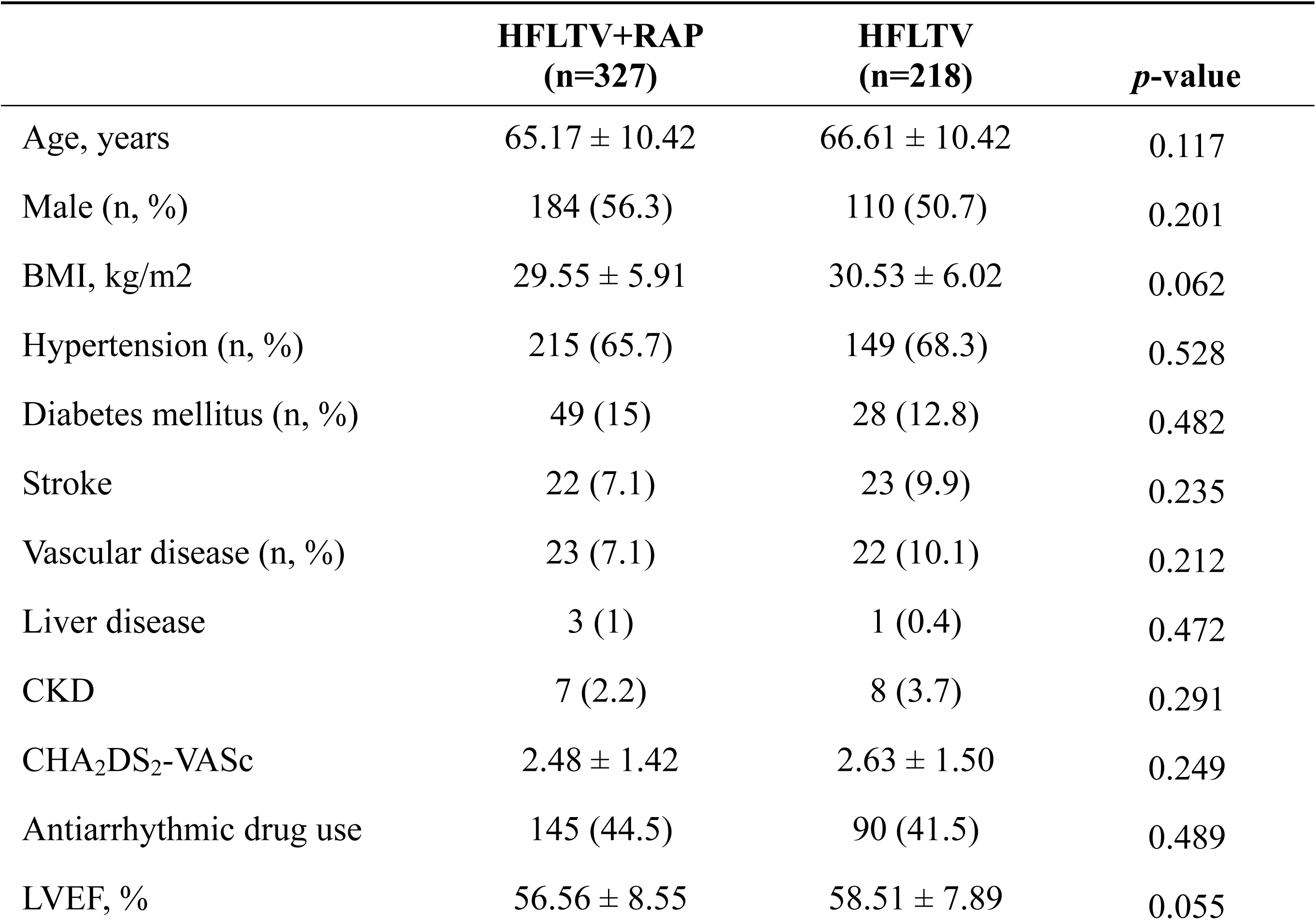
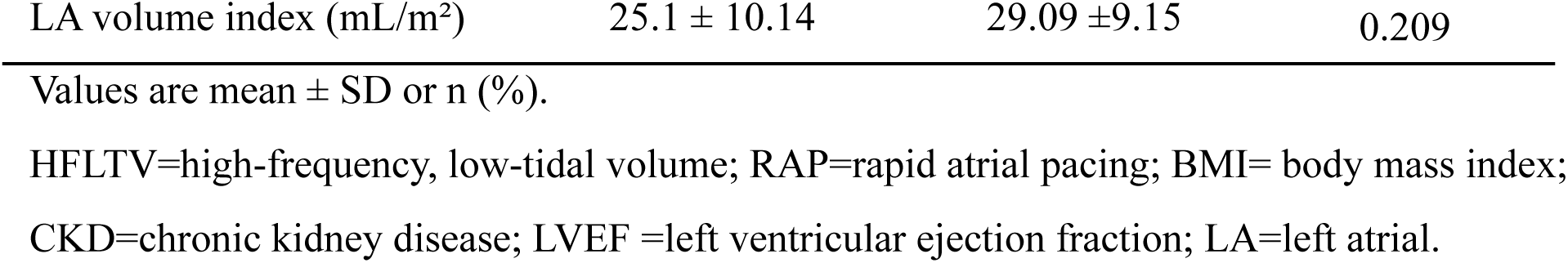
Baseline characteristics.

### Procedural Efficiency

There were no statistically significant differences in procedural (HFLTV+RAP 74 min [57-98] vs. HFLTV 66 min [IQR 53-85.75], p=0.617) and total RF ablation times (HFLTV+RAP 15.15 min [11.22-21.22] vs. HFLTV 13.99 min [11.04-17.13], p=0.620) between the groups (**Table 2**, **Figure 2**).

**Figure 2.**
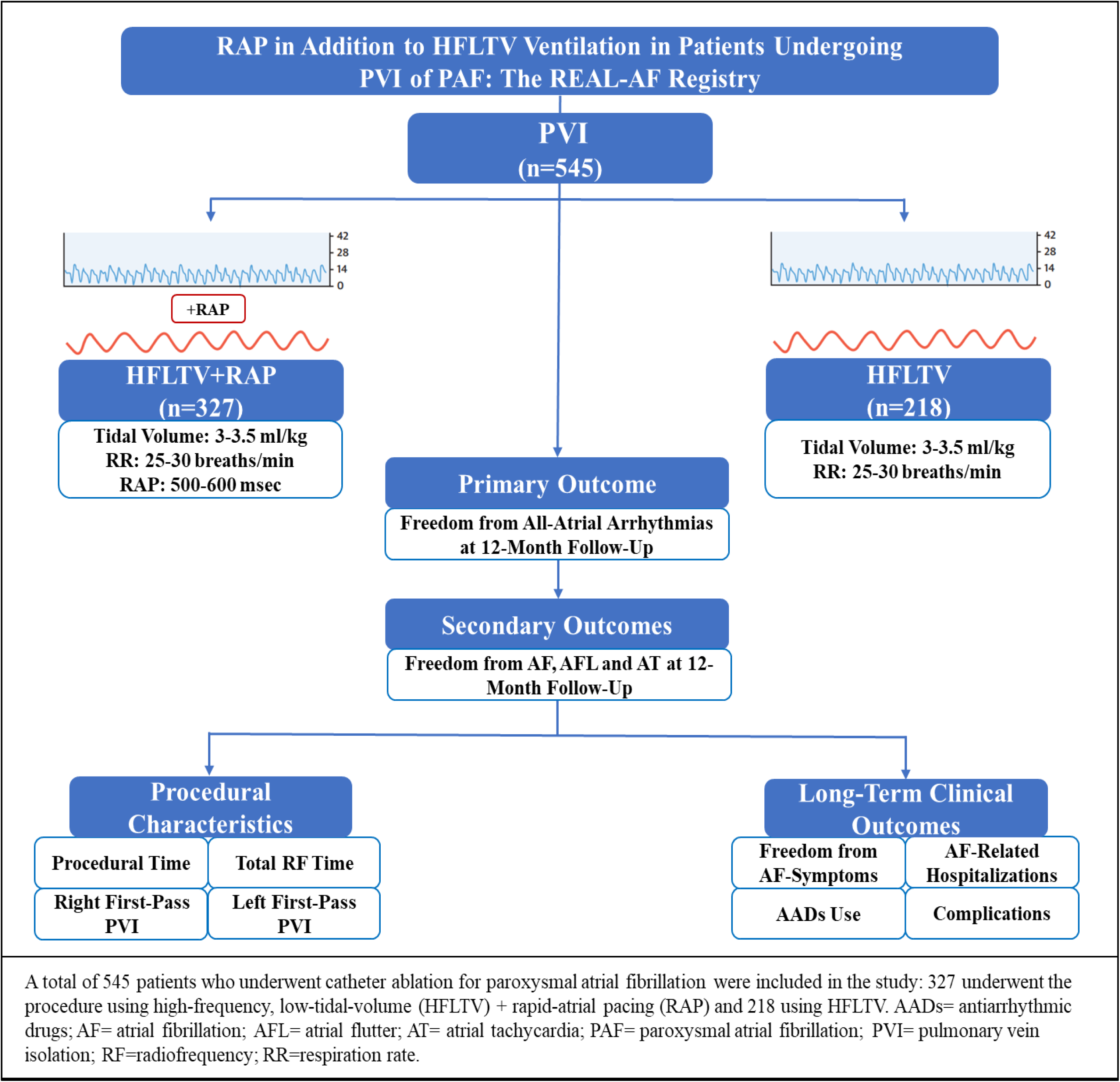
Procedural characteristics. A) Procedural time. B) Total RF time. C) First-pass PVI. Abbreviations as in Figure 1.

**Table 2.**
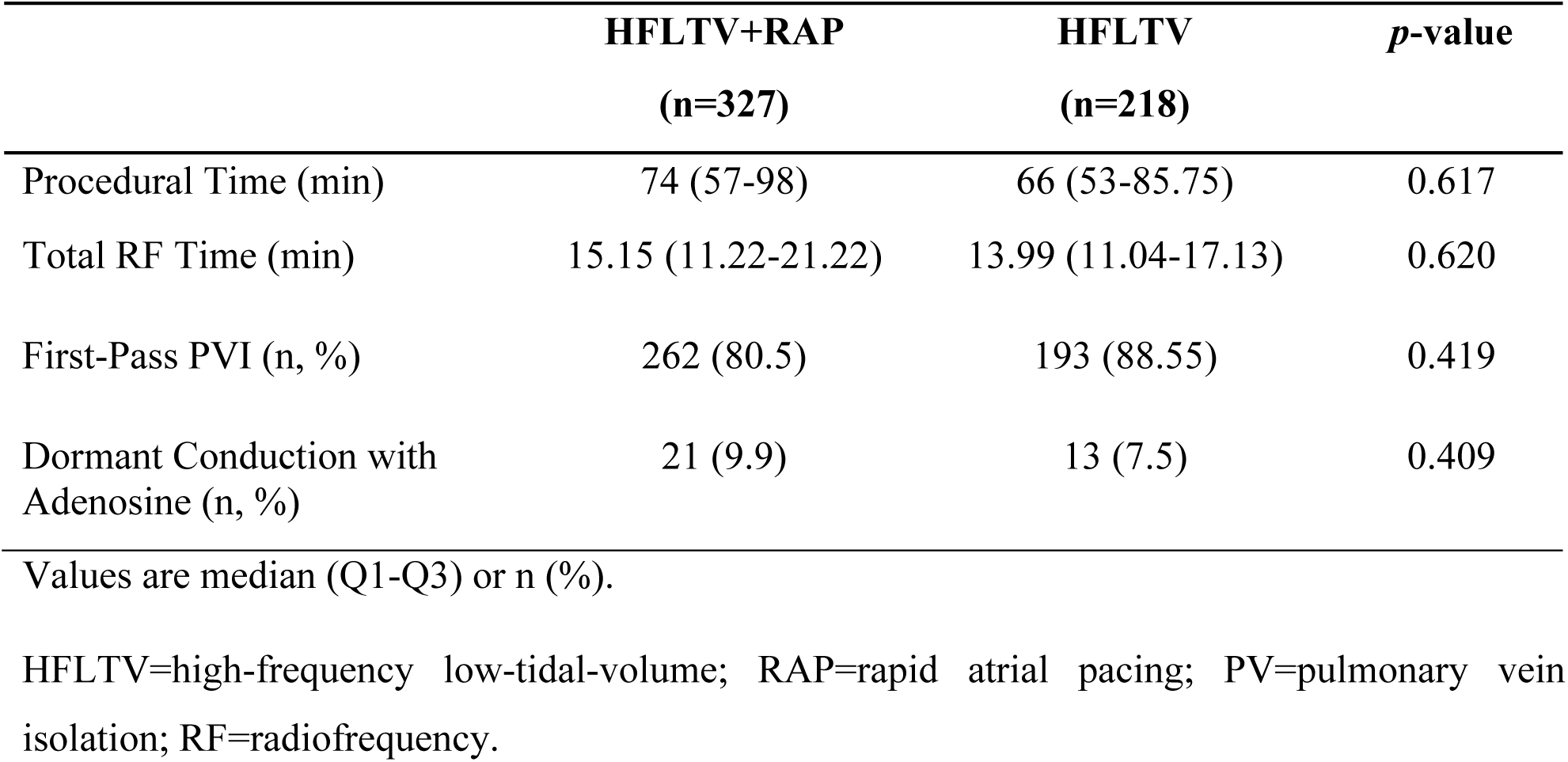
Procedural characteristics.

|  | **HFLTV+RAP**<br>**(n=327)** | **HFLTV**<br>**(n=218)** | ***p*-value** |
| --- | --- | --- | --- |
| Procedural Time (min) | 74 (57-98) | 66 (53-85.75) | 0.617 |
| Total RF Time (min) | 15.15 (11.22-21.22) | 13.99 (11.04-17.13) | 0.620 |
| First-Pass PVI (n, %) | 262 (80.5) | 193 (88.55) | 0.419 |
| Dormant Conduction with Adenosine (n, %) | 21 (9.9) | 13 (7.5) | 0.409 |

Values are median (Q1-Q3) or n (%).
HFLTV=high-frequency low-tidal-volume; RAP=rapid atrial pacing; PV=pulmonary vein isolation; RF=radiofrequency.

### Acute Procedural Efficacy

The rate of first pass PVI was similar within both groups (HFLTV+RAP 80.5% vs. HFLTV 88.55%, p=0.419) (**Figure 2**). The majority of RFCA procedures were performed with zero or minimal fluoroscopy. There was no difference in the rate of unmasking dormant conduction with adenosine between the groups (HFLTV+RAP 9.9% vs. HFLTV 7.5%, p=0.409) (**Table 2**).

### Primary Outcome

The mean follow-up duration was 369.7 ± 78.9 days, and 356 ± 95.7 days, respectively for HFLTV+RAP and HFLTV ventilation populations (p=0.053). There was no statistically significant difference in freedom from all-atrial arrhythmias at 12 months of follow-up between the groups (HFLTV+RAP 82.68% vs. HFLTV 86.52%, HR=1.43, 95% CI (0.94-2.16), p=0.093) (**Figure 3**).

**Figure 3.**
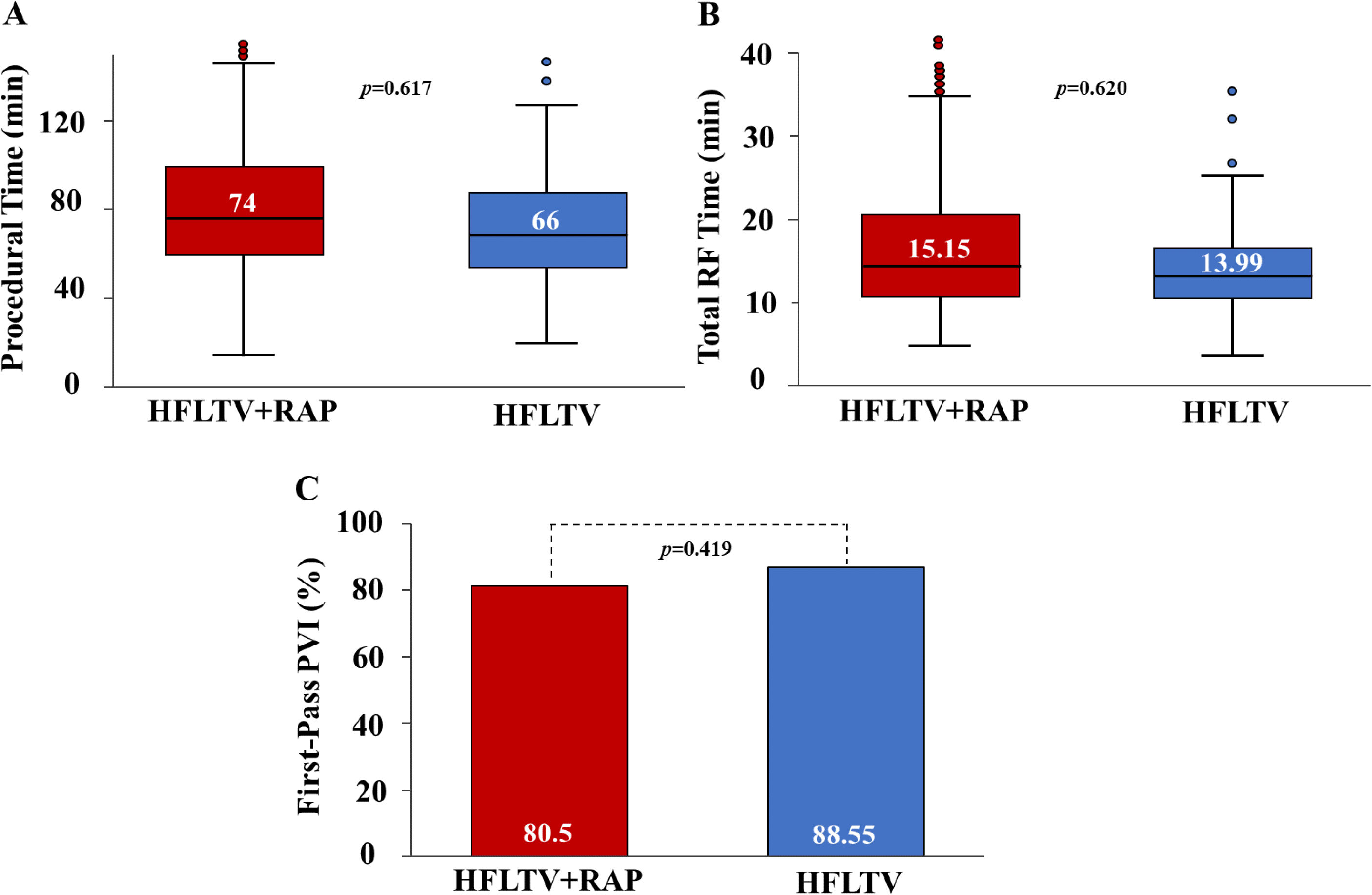
Freedom from all-atrial arrhythmias at 12 months of follow-up. Abbreviations as in Figure 1.

### Secondary Outcomes

The analyses for each subtype of atrial arrhythmia (AF, AFL, and AT) are displayed in **Figure 4**. Freedom from AF at 12 months of follow-up was significantly lower in the HFLTV+RAP group compared to the HFLTV ventilation alone group (HFLTV+RAP 84.6% vs. HFLTV 87.67% HR=1.45, 95% CI (1.02-2.07), p=0.039). There were no statistically significant differences in freedom from AFL (HFLTV+RAP 96.16% vs. HFLTV 97%, HR=1.59, 95% CI (0.36-7.04), p=0.5) and AT (HFLTV+RAP 97% vs. HFLTV 98%, HR=1.09, 95% CI (0.30-4.03), p=0.9) at 12-months of follow-up (**Figure 4**).

**Figure 4.**
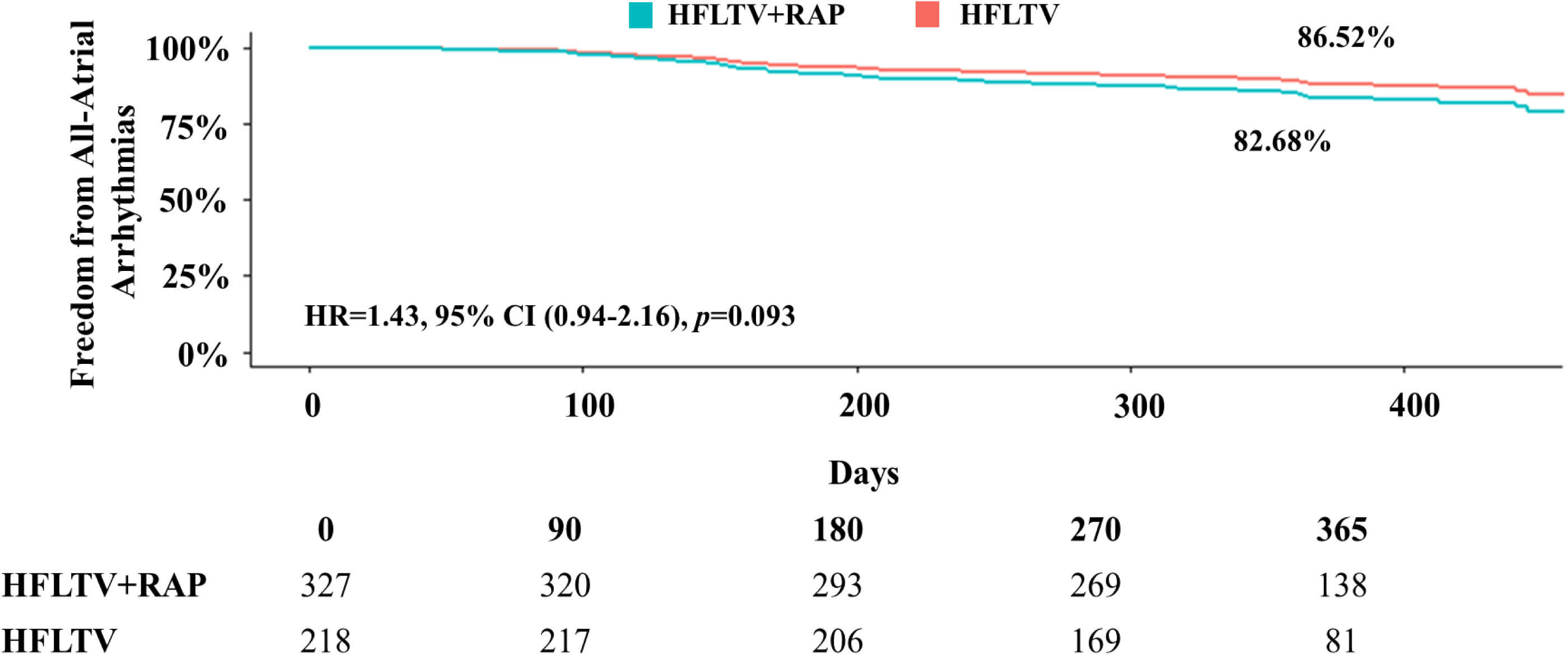
Freedom from each subtype of atrial arrhythmia at 12-months of follow-up. A) Atrial fibrillation. B) Atrial flutter. C) Atrial tachycardia. Abbreviations as in Figure 1.

### Long-Term Clinical Outcomes

The freedom from AF-related symptoms was similar between the groups (HFLTV+RAP 91.4% vs. HFLTV 93.1%, p=0.476). Furthermore, the rate of AF-related hospitalizations was comparable (HFLTV+RAP 1.5% vs. HFLTV 2.8%, p=0.320) within both groups. There was no significant difference in the use of AADs at 12 months of follow-up (HFLTV+RAP 4% vs. HFLTV 4.6%, p=0.728) (**Figure 5**).

**Figure 5.**
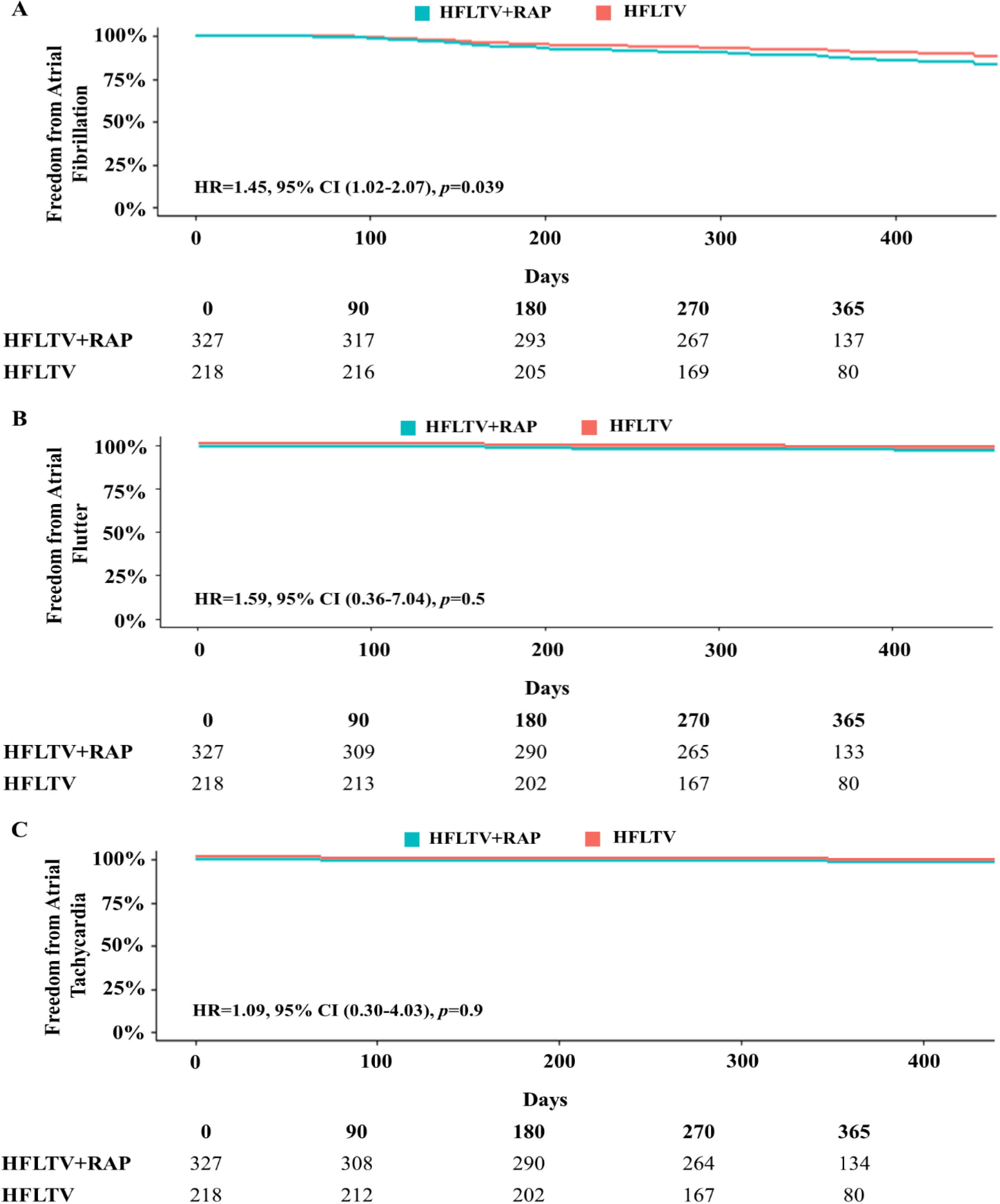
Long-term clinical outcomes at 12 months of follow-up. A) Freedom from AF-related symptoms. B) Rate of AF-related hospitalizations. C) AADs use. D) Procedure-related complications. Abbreviations as in Figure 1.

### Safety Outcomes

All patients tolerated the HFLTV ventilation protocol without anesthesia-related complications. There were no clinically relevant events of CO_2_ retention. There were no significant differences in acute (HFLTV+RAP 0.3% vs. HFLTV 0%, p=0.414) and long-term procedure-related complications (HFLTV+RAP 0.6% vs. HFLTV 0%, p=0.247) within the groups. One complication corresponded to a vascular access adverse event (groin hematoma), and the other was a patient who presented with post-procedural pericarditis. Both complications occurred in the HFLTV+RAP group and were managed clinically with no need for surgical intervention. All patients in the HFLTV+RAP group tolerated the RAP protocol with no pacing-associated complications. No patients required prolonged mechanical ventilation or reintubation after the procedure (**Table 3**).

**Table 3.** Acute and long-term complications.

| Type of Complication | HFLTV+RAP<br>(n=327) | HFLTV<br>(n=218) | *p*-value |
| --- | --- | --- | --- |
| Stroke | 0 | 0 | 1.00 |
| Transient Ischemic Attack | 0 | 0 | 1.00 |
| Pericardial Effusion/<br>Cardiac Tamponade | 0 | 0 | 1.00 |
| Phrenic Nerve Injury | 0 | 0 | 1.00 |
| Atrioesophageal Fistula | 0 | 0 | 1.00 |
| Pericarditis | 1 (0.3) | 0 | 0.389 |
| Groin Hematoma | 1 (0.3) | 0 | 0.414 |

Values are n (%).
HFLTV=high-frequency low-tidal-volume; RAP=rapid atrial pacing.

## Discussion

To the best of our knowledge, this is the first prospective, multicenter study comparing procedural and long-term clinical outcomes of RFCA for PAF with HFLTV ventilation plus RAP versus HFLTV ventilation alone. Our principal findings were as follows:

1. RAP was not associated with any additional benefit to HFLTV ventilation in terms of procedural efficiency and efficacy (procedural and RF times, first pass PVI).
2. RAP in addition to HFLTV ventilation, was not associated with an improved freedom from all-atrial arrhythmias at 12-months of follow-up compared with HFLTV ventilation alone.
3. Combining HFLTV ventilation with RAP was not associated with better long-term clinical outcomes (freedom from AF-related symptoms, rate of AF-related hospitalizations, and AADs use) at 12-months of follow-up compared to HFLTV ventilation alone.

HFLTV ventilation has been demonstrated to improve procedural and long-term clinical outcomes in RFCA of PAF and PeAF compared to SV, with a similar safety profile (24,25). Previous small RCTs have shown that RAP improves procedural characteristics during RFCA of PAF, particularly catheter stability, thus enhancing ablation lesion quality (21,26). Furthermore, especially in bradycardic patients, RAP may not only reduce stroke volume due to shorter diastolic filling time, but it may increase the rate of data acquisition during mapping. However, the potential long-term clinical benefits of incorporating RAP in conjunction with other ventilation strategies during RFCA of PAF had not yet been studied. Our findings indicated that adding RAP to HFLTV ventilation was not associated with a significant improvement in procedural or long-term clinical outcomes. Conversely, RAP showed a trend for longer procedural times. Although this difference was not statistically significant, this finding highlights important considerations for clinical practice. The routine addition of a CS catheter for RAP, while theoretically beneficial during electrophysiology studies and in scenarios involving persistent or longstanding PeAF, where the induction of non-PV triggers and mappable arrhythmias such as atrial tachycardia, typical and atypical AFL, is more frequent, may not be necessary for RFCA of PAF, particularly if PVI is the only lesion set delivered. Therefore, the placement of a CS catheter during PAF ablation, which may require obtaining additional femoral access depending on the operator’s practice, could be considered redundant. This could potentially increase procedural complexity, time, and complication risk without offering substantial clinical benefit. Longer anesthesia times, the necessity for extra vascular access, the risk of CS laceration or perforation, are all considerations that may weigh against the routine use of RAP in RFCA of PAF. Importantly, a recent study demonstrated that CS perforation is responsible for 7% of all cases of myocardial perforation requiring open heart surgery (31). Moreover, it may also increase fluoroscopy times, particularly in operators who do not perform fluoroless RFCA.

Conversely, it appears that performing RAP at 500-600 milliseconds does not improve catheter stability during RFCA of PAF when HFLTV ventilation is employed. Cardiac atrial pacing rate was arbitrarily set based on the assumption of patient tolerability, but it is unknown if a different pacing rate could provide further benefits. There is a lack of evidence about whether there is a lower rate threshold in bradycardic patients for starting RAP to improve outcomes. Prior RCTs have evaluated procedural outcomes with RAP (21,26), nevertheless, our methodology differed significantly from that of the previously mentioned RCTs. These studies involved small sample sizes (20-40 patients) and utilized low-power, long-duration (LPLD) ablation settings (30W, 30 seconds per lesion). In contrast, our cohort comprised a much larger sample size (545 patients), and all RFCA procedures were performed using HPSD ablation, which has already demonstrated a positive impact on procedural outcomes in AF ablation (14). In addition, while Aizer at el. (21,26) compared procedural outcomes with RAP during the first half versus the second half of lesion creation, we implemented continuous RAP throughout the entire ablation procedure and in combination with HFLTV ventilation. Although these RCTs indicated that RAP application reduced CF variability, thereby improving catheter stability; our findings suggested that this improvement was not necessarily associated with significant procedural, short- and long-term clinical benefits. Besides, the use of RAP may entail higher costs due to the potential necessity of an additional vascular access, and the requirement for a mapping CS catheter, which can considerably increase the expenses associated with a routine procedure such as PAF ablation.

HFLTV ventilation has demonstrated to improve catheter stability by minimizing diaphragmatic excursion, thereby influencing lesion transmurality. Likewise, RAP has shown to also enhance catheter stability and lesion formation by inducing heart rate acceleration. Although, these techniques may not completely eliminate thoracic excursion. Our results exhibited that the impact of diaphragmatic movement on CF variability may outweigh that of cardiac motion. Hence, the implementation of HFLTV ventilation may obviate the need for the additional stability benefits provided by RAP (32). Besides, it is relevant to recognize that success rates can be also influenced by additional factors beyond thoracic excursion, underscoring the importance of identifying other ablation parameters that reliably predict lesion quality (33).

Simultaneous pacing during AF ablation has not demonstrated improvement in safety outcomes by mitigating adverse events associated with elevated CF measurements, encompassing steam pops, thrombus formation, and cardiac perforation (21,26). Our study showed no significant differences in procedure-related complications between the groups. While RAP has been proven to have an adequate safety profile when used in conjunction with HFJV, it does not offer any additional safety benefits when used with HFLTV ventilation (34). In summary, while RAP in conjunction with HFLTV ventilation may have theoretical and practical applications in more complex AF ablation scenarios, our study suggested that its routine use in PAF ablation was not associated with significant procedural or long-term clinical benefits. Future research should focus on identifying specific patient subsets or procedural contexts where RAP may indeed offer a tangible advantage, thus refining ablation strategies for optimal patient outcomes.

Finally, it is important to recognize that some operators simply exchange the intracardiac echocardiography (ICE) catheter for a multispline catheter and perform RAP from the SVC. Although this approach eliminates the need for additional femoral venous access or catheters, it may forfeit the potential advantages of ICE, such as real-time lesion formation monitoring and continuous assessment of pericardial effusion throughout the procedure.

## Study Limitations

First, this was an observational study; although baseline clinical characteristics were comparable between the groups and participants were prospectively enrolled, unmeasured confounders might still be present, opening the possibility for selection bias. Notably, our study incorporated clustering Cox Proportional Hazards models to account for individual operator effects, allowing a fair comparison between highly experienced and less experienced operators while also controlling for variations across different centers. Ultimately, cardiac pacing was set to 500-600 milliseconds in the HFLTV+RAP group based on assumed patient tolerability; however, it remains unclear if a different pacing rate could provide any further benefits.

## Conclusion

In patients undergoing RFCA for PAF, the addition of RAP to HFLTV ventilation was not associated with improved procedural and long-term clinical outcomes at 12-month follow-up. Therefore, the role of placing additional catheters to perform RAP during RFCA of PAF should be further investigated.

### Funding and Disclosures

The REAL-AF registry is funded through an investigator-initiated research grant (Dr. Osorio, principal investigator) from Biosense Webster. Dr. Osorio, Dr. Zei, Dr. Sauer, and Dr. Romero report consulting and research support from Biosense Webster. Dr. Silverstein reports consulting and honoraria from Biosense Webster. Dr. D’Souza and Dr. Rajendra report consulting from Biosense Webster. Dr. Oza reports advisory board and physician education from Biosense Webster. Dr. Thosani reports physician education from Biosense Webster. The other authors have reported no relationships relevant to the contents of this paper to disclose.

## Abbreviations

HFLTV: High-frequency low-tidal volume
HFJV: High-frequency jet ventilation
HPSD: High-power short-duration
LVEF: Left ventricular ejection fraction
PAF: Paroxysmal atrial fibrillation
PVs: Pulmonary vein/s
PVI: Pulmonary vein isolation
RFCA: Radiofrequency catheter ablation
RAP: Rapid atrial pacing
SV: Standard ventilation

## Data Availability

All the data referred to in the manuscript is available.

## Notes

### Clinical Trial

NCT04088071

### Author Declarations

The REAL-AF registry was approved by the Western Institutional Review Board?Copernicus Group on October 31, 2018. Furthermore, institutional review boards of each participating institution approved the participation of enrolling centers adhering to the principles outlined in the Declaration of Helsinki.

